# ASSESSING HOME DELIVERY MODEL OPTION FOR IMPROVING ACCESS TO ANTIRETROVIRAL THERAPY; PROTOCOL FOR A CLUSTER- RANDOMIZED TRIAL IN ANAMBRA STATE, NIGERIA

**DOI:** 10.1101/2021.04.27.21256089

**Authors:** Ajagu Nnenna, Offu Ogochukwu, Nduka Sunday, Ekwunife Ikechukwu Obinna

**Affiliations:** Department of Clinical Pharmacy and Pharmacy Management, Nnamdi Azikiwe University, Awka, Nigeria; Department of Clinical Pharmacy and Biopharmaceutics, Enugu State University of Science and Technology, Agbani, Nigeria

**Author notes:** Corresponding author: Ajagu Nnenna, Department of Clinical Pharmacy and Pharmacy Management, Nnamdi Azikiwe University, Awka, Nigeria and Department of Clinical Pharmacy and Biopharmaceutics, Enugu State University of Science and Technology, Agbani, Nigeria. Author information Ajagu Nnenna (Msc.), Offu Ogochukwu (Msc.), Nduka Sunday (PhD), Ekwunife Obinna (PhD).

**Keywords:** Antiretroviral, HIV, Home delivery, Nigeria

## Abstract

**Background:** Although different antiretroviral (ARV) agents’ delivery models have been explored in Nigeria, yet only about 55% and 35% of affected adults and children respectively are currently on these agents. This, therefore, underscores the need for the identification of newer effective antiretroviral therapy delivery models and best strategies to increase access to ARV therapy in Nigeria.

**Method and Analysis:** The study will be a cluster randomized controlled trial, which will be conducted in two HIV treatment centers in Anambra State, Nigeria. Participants will be randomized into intervention and control arm. Home delivery personnel will be trained to deliver the ARV drugs at 3months intervals to the homes of those in the intervention group while those in the control group (Facility-based services group) will receive ARVs at the HIV treatment hospital. The primary outcome for the trial will be the difference between groups in the proportion with HIV viral load suppression (≤20 copies/mL) by 12 months and then 24months. Secondary outcome include adherence to ARVs, the cost-effectiveness of home delivery service, and patient satisfaction.

**Ethics and dissemination:** Nnamdi Azikiwe University Teaching Hospital Health Research Ethics Committee (NAUTHHREC) approved this study (NAUTH/CS/66/VOL.13/VER III/23/2020/011) with a trial registration: Pan African Clinical Trials Registry, ID: PACTR202004535536808 on 8th April 2020. Study dissemination plans include holding advocacy meetings with stakeholders in HIV care, publishing study outcomes in an open-access peer-review journal, and presentation of the outcome at an international conference.

*Strength and Limitations of this study:*

- This is the first randomized trial assessing the effectiveness of the home delivery model of antiretroviral therapy.
- The primary outcome in this trial will be measured by assessing the difference between baseline HIV viral load and 12/24 months in both the intervention and control arm.
- In this study, patients will be assigned to treatment groups and non-treatment groups. This will help us to compare the effect of the home delivery model.
- In this study, only one state in Nigeria and two HIV/AIDS treatment hospitals in the state will be used to access the impact of the home delivery model. This may not allow for the generalization of the study findings.
- This study could serve as a pilot to a full-fledged national study to access the effectiveness of the home delivery model.

## INTRODUCTION

We owe approximately 39 million people who have died of HIV/AIDS the significant obligation to eradicate the disease. Additionally, eradication of this disease represents a tremendous opportunity to lay the foundation for a healthy world for the future generation (1). Unfortunately, the morbidity and mortality rate from HIV/AIDS is still high (2) due to some important reasons including poor access to highly active antiretroviral therapy (HAART) (2).

Poor access and adherence to the HAART has a significant impact on the overall treatment outcomes because of the chronic nature of the disease (3). Hence, continuous access to HIV treatment is very important to achieving a successful positive response to this public health issue (2).

It is the right of everyone including people living with HIV to have equitable access to the drugs as contained in sustainable development goals (SDGs) (4), and the right to equitable health is linked to one of the targets set by the Joint United Nations Programme on HIV and AIDS (UNAIDS) for ending HIV in the year 2030. According to this target, at least 95% of people living with HIV should have access to antiretroviral agents (5) as this is strategic to achieving the clinical, immunologic, and virologic goals of HIV/AIDS treatment. Ultimately, ending HIV can only be achieved when the right to access drugs is placed at the center of the public health campaign to end HIV (6).

About 36.9 million people are living with HIV/AIDS globally and only about 59% of this population had access to ARV according to the 2019 UNAIDS factsheet (7). Whereas Africa constitutes about 67% of the global disease burden(8) and with about 1.9 million of them living in Nigeria with a prevalence rate of 1.4 %(9), only 55% and 35% of the affected adults and children respectively had access to ART in 2018 (10). Hence, the identification of new strategies to increase access to ARV agents should be an urgent priority (11).

In 2016, the World Health Organization (WHO) modified its CD4 cell count threshold in the treatment guidelines and recommended a test-and-treat strategy for all persons living with HIV (12). This development substantially increased the number of people on ART (13) with an increasing cohort of stable patients which may constitute a huge burden to the already burdened health system, especially in most developing countries. Therefore the development of new and innovative ways to increase access to antiretroviral therapy that would decongest health facilities while supporting the SDG goals is of utmost importance.

A semi-structured interview carried out by Hardon et al in 2007(14) in health facilities in Uganda, Tanzania and Botswana identified convenience, long waiting times to receive care, and transportation cost as challenges to achieving optimal care in HIV/AIDS management. A study by Cunningham et al., (15) in the US, reported the cost of transport to the treatment site and the inability to get out of work as important barriers to accessing ART. Similarly in Nigeria, long distances to the service delivery points, transport costs, as well as long waiting times have been identified as barriers to accessing antiretroviral agents in the country (16). Meanwhile, a study by Jaffar et al. (11) that compared rates of virologic failures in patients treated at home as against those treated in the facility (hospital) found no significant difference between the two.

Different strategies to improve access to ARVs in Nigeria have been suggested by the Strengthening Integrated Delivery of Services on HIV/AIDS (SIDSHAS) projects and currently in their pilot stage. Among these strategies are the Community Antiretroviral Refill Club (CARC) for patients living in remotes areas with a particular interest in the riverine areas and the Community Pharmacy Antiretroviral Refill Programme (C-PARP)(17). These programs are targeted at reducing visits to the hospitals by stable clients to decongest the facilities which come with several other benefits.

Meanwhile, a theoretical foundation underlines the delivery of antiretroviral to patients’ homes as a way of improving both adherence and viral load, as seen in the HUB and SPOKES model of differentiated care, where centers provide specialized services through established linkages with several secondary facilities within a network and patients are referred to for specialized services when the indications for such arises (18–21). Services provided at this level include HCT, ARV re-fill, adherence counseling, and treatment of simple opportunistic infections (OIs) (18–21). This could serve as a current focus strategy for improving access to ARVs. The practical benefit of this approach is a significant reduction in the pharmacy waiting time while decongesting the healthcare facilities. This approach has been successfully applied in some facilities in the UK (22) and hence, it looks like a promising strategy to achieving improved access to ARVs in resource-limited settings. This study, therefore, aims to assess the possibility of using a home-based antiretroviral delivery model and as a new strategy for increasing access to antiretroviral agents in resource-limited settings.

## METHODS AND ANALYSIS

### Study design

This study will utilize a cluster-randomized trial approach to evaluate the effectiveness of antiretroviral home delivery strategy on HIV/AIDS treatment outcomes with a focus on improving access to the ARV agents. Based on the information we received at the time of conducting this study, only 12 hospitals in Anambra State offer comprehensive HIV services including HIV-adherence counseling and antiretroviral treatment services. Two hospitals among the 12 comprehensive HIV treatment hospitals that have high HIV/AIDS patients’ base were randomly selected. Patients that meet the inclusion criteria in these two hospitals will be randomly assigned to intervention or control (see fig 1).

**Fig 1.**
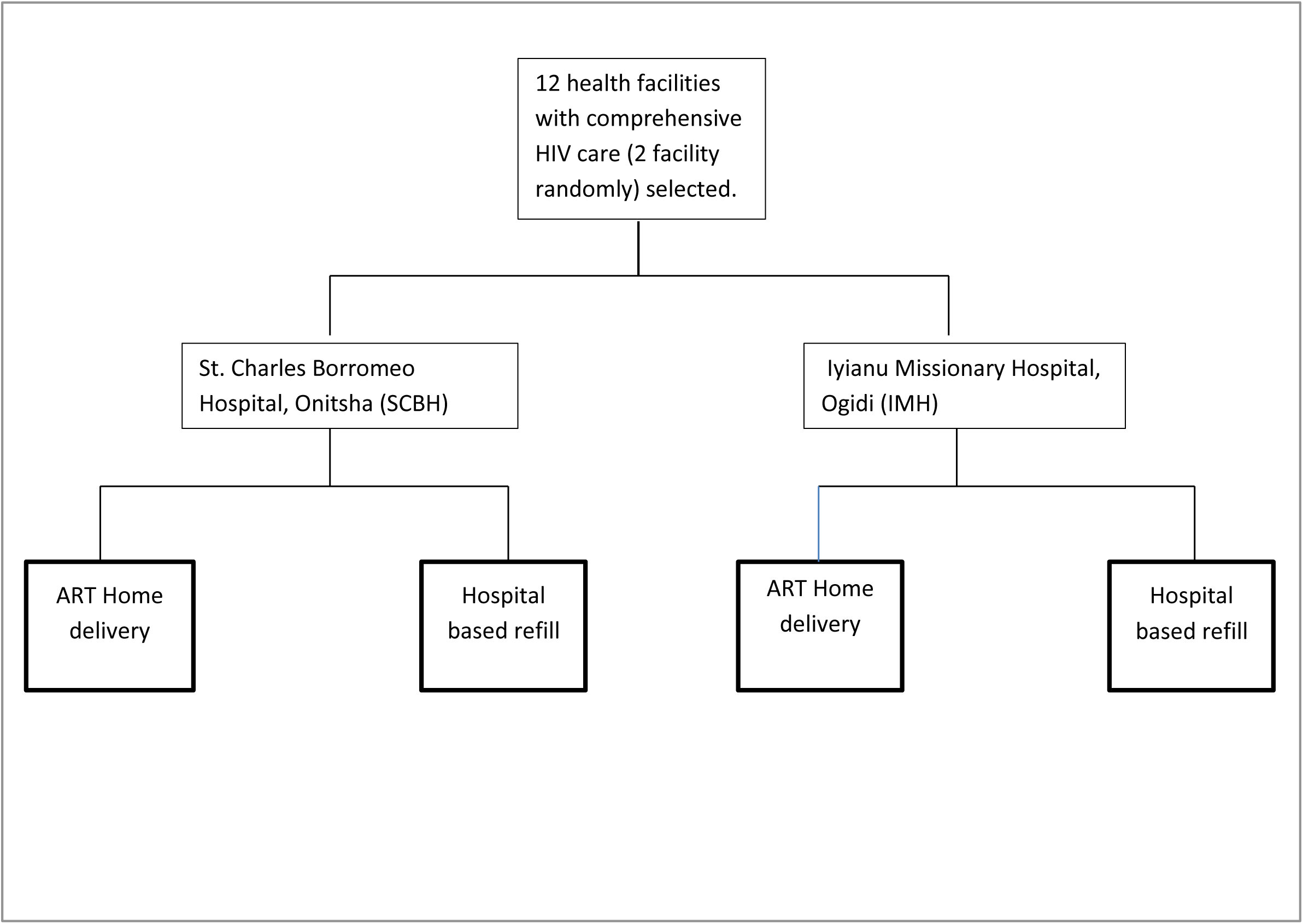
Randomization Outline

**Fig 2.**
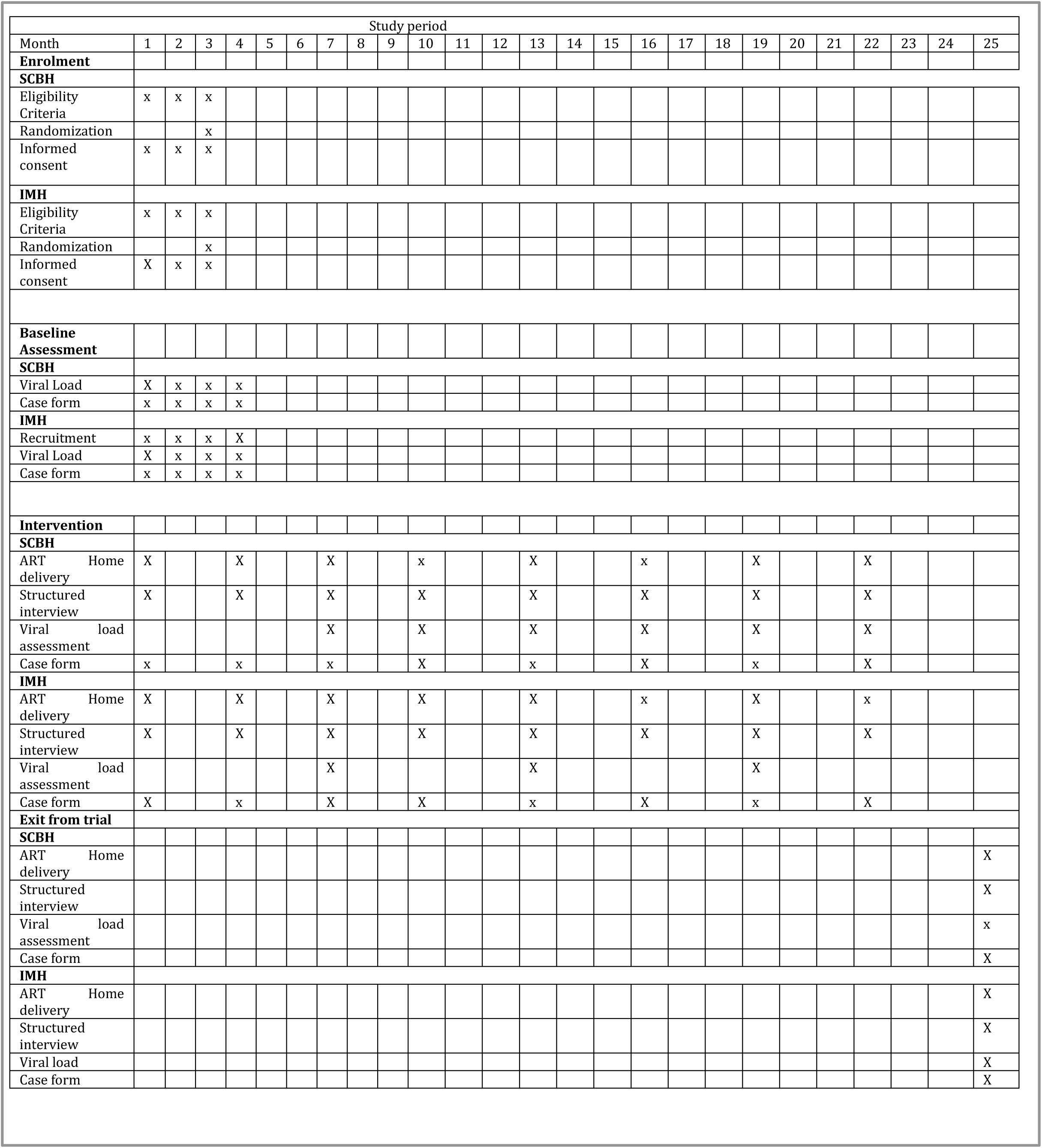
Trial Schedule

### Study setting

The study setting will be in Anambra state which is one of the five states in the Southeast geopolitical zone in Nigeria. It is bounded in the northeast by Enugu state, in the east by Enugu and Abia states, in the west by Delta state while in the South and Northwest by Imo and Kogi states respectively. The State is mainly inhabited by Igbo-speaking people who are mostly Christians. Most members of the population are farmers, civil servants, artisans, and businessmen, and women (23); with a projected population of approximately 4,177,828 (50.7% males and 49.3% females) (24). Interestingly, about 60.5% of these populations are of ages 15-65 years (25). Awka is the state’s capital and there are 21 local government areas in the state out of which five are urban; five are semi-urban while eleven are rural (23). Anambra state is made up of 21 local government areas with an HIV prevalence rate of 2.4% (25). There are about 1,485 health facilities in the state including two tertiary and 31 public health facilities, with private hospitals constituting about 75% of the health facilities (26).

### Intervention

Enrolled participants will be directed to the research assistant responsible for introducing the study to the enrollees and they will consent to the study by signing the consent form (see Fig 3). Those that give their consent will be sent to the physician after collecting information that will aid home delivery of their drugs including delivery points, preferred day of the week, and most available phone numbers among others. The physician will then, examine the patients and request a baseline viral load.

**Fig 3:**
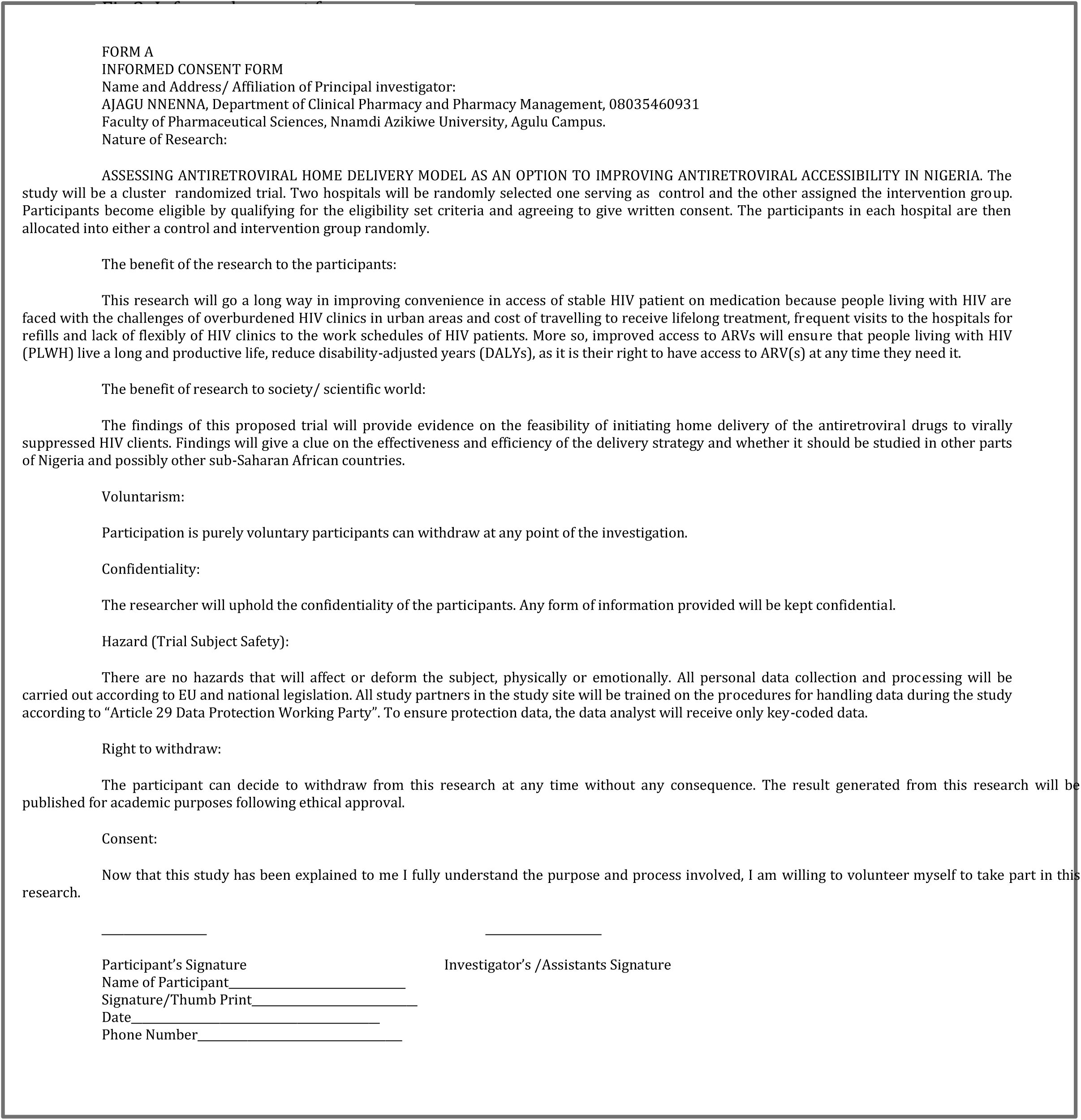
Informed consent form

Based on the physician’s examination and findings, the Home Delivery Personnel (HDP) will deliver a three-month prepackaged ARV in a suitable package form with a patient identifier to the patient’s chosen delivery point using a delivery motorcycle. The delivery points are chosen and the drugs are delivered in a way to ensure confidentiality. Patients in this arm will be advised to visit the hospital every six months for viral load and CD4+ cell count unless there is a problem requiring the physician’s attention. Meanwhile, the HDP will be trained on how to interview these patients during the drug delivery using short structured interview questions (see Fig. 4) while monitoring for adherence. The HDPs after each delivery reports back to the physician who then decides on whether to schedule the patient on appointment. Participants in this arm will be provided with a phone number to call in case of emergency or urgent consultation needs before the next appointment schedule.

**Fig. 4.**
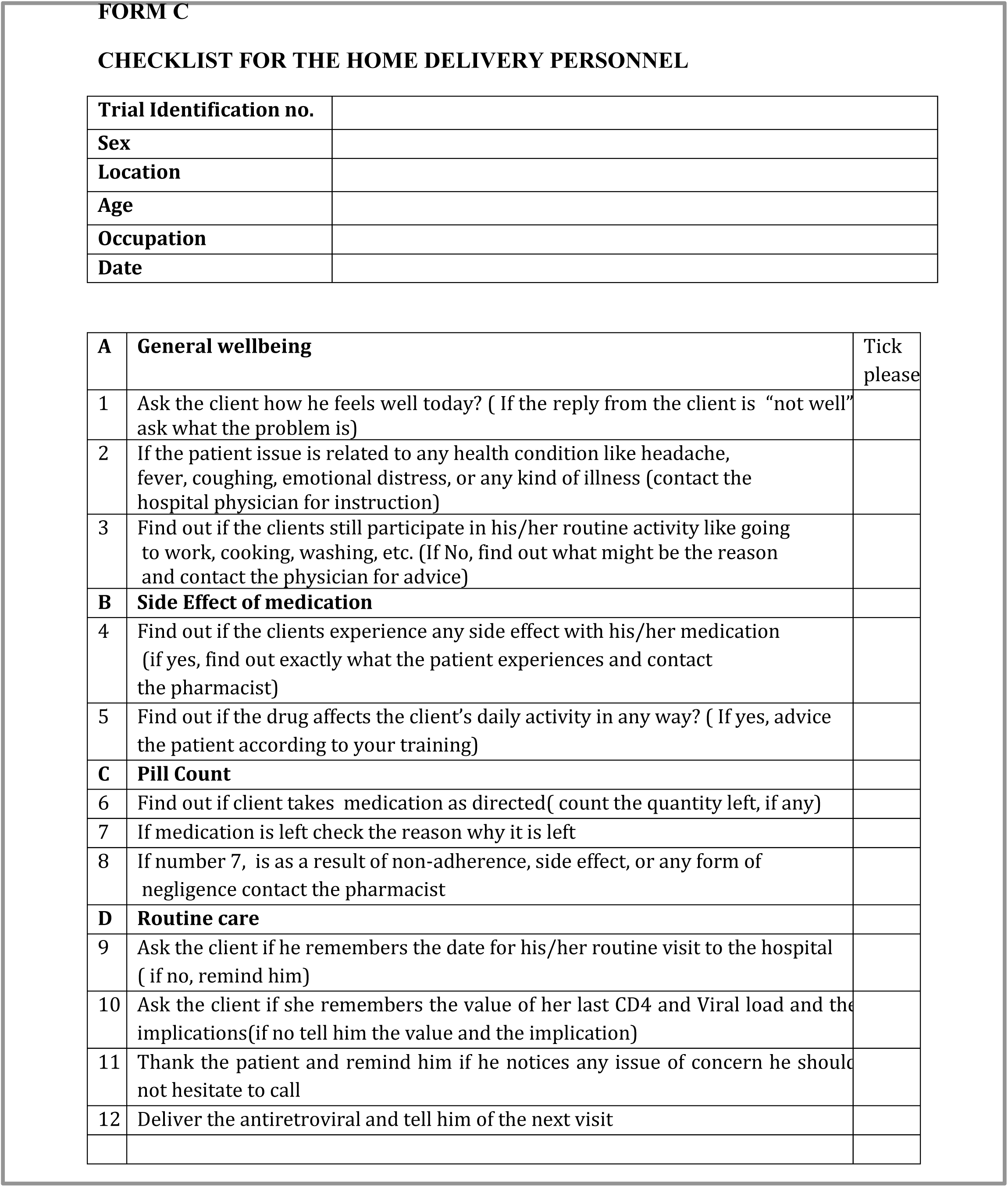
checklist for the home delivery personnel.

**Fig 5:**
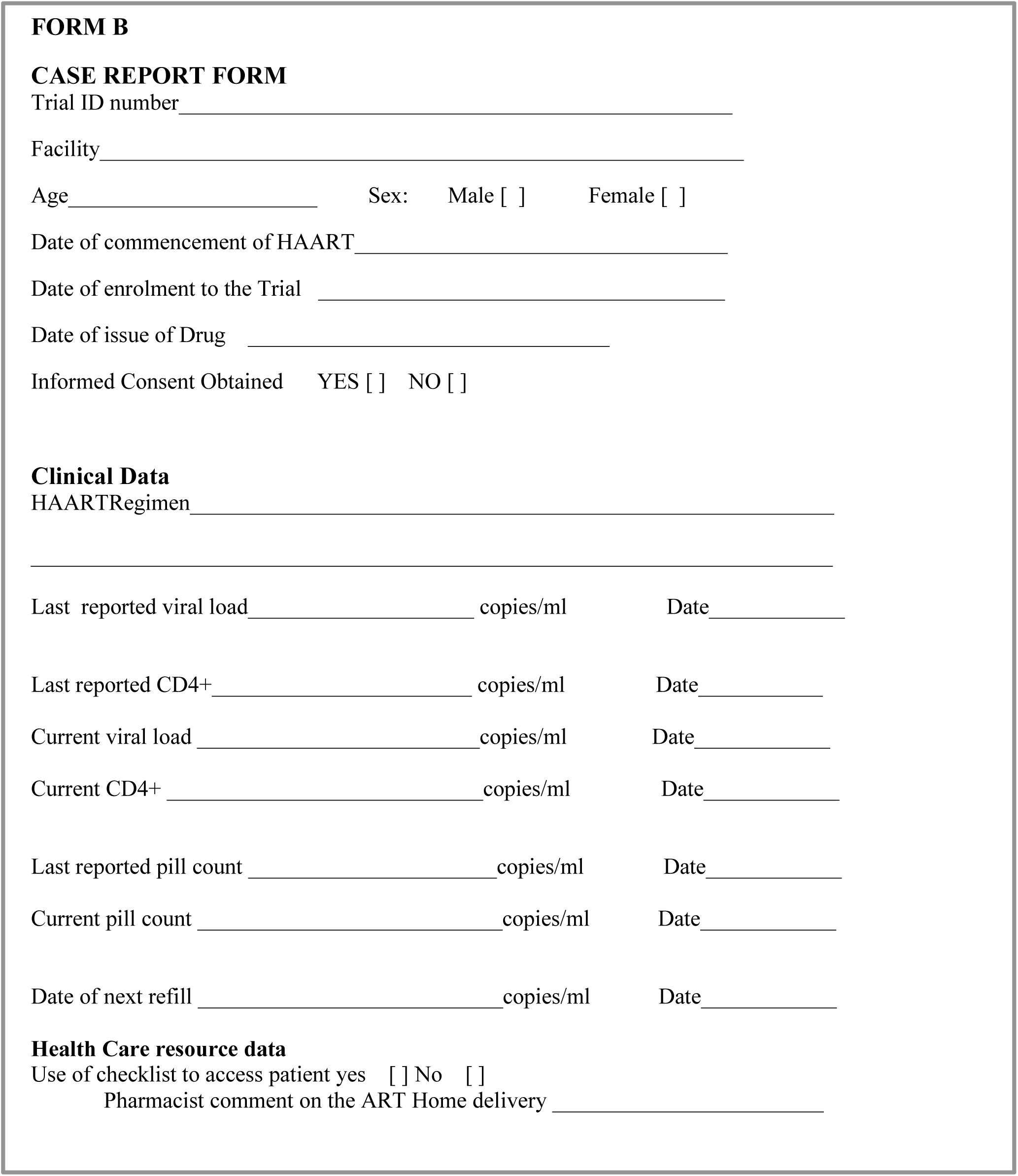
Case Form

On the other hand, the control group will receive their drugs in the HIV treatment hospital as usual but assess the viral load and CD4 count every six months as in the intervention group. Routine data will be collected and the patient is at liberty to withdraw from the trial at any time.

### Home delivery personnel training

The community health extension worker (CHEW) who will serve as the Home delivery Personnel (HDP) will undergo a 5-day training using the National ART training curriculum, World Health Organization (WHO) guideline on management of stigma, and WHO community home delivery manual. The HDPs were also trained on how to maintain utmost confidentiality when delivering the medication. The training will be carried out by an expert on HIV services.

### Eligibility criteria

#### Inclusion criteria

All Stable patients who commenced ART for ≥ 6months and have suppressed viral load.

#### Exclusion criteria

i. Pregnancy at the time of enrollment
ii. Age < 18years
iii. Defaulters
iv. Unsuppressed viral load (>20 copies /mil)
v. Withdrawal of consent
vi. Reside outside the state

### Sample Size

Using a web-based sample size calculator,(27) and to demonstrate an HIV viral load suppression difference of ≤20 copies/ml between the control and the intervention arm between the baseline and 12 months and 24 months with a one-sided p-value of 0.05 and a power of 80%, a minimal sample size of 264 patients was deemed appropriate for each study group. To account for possible dropouts and loss to follow-up, an extra 20% of the calculated sample will be added to bring the number to a total of 316 patients of 158 patients per group.

### Recruitment and retention

HIV patients that are eligible for the study will be recruited by the study personnel in the trial site (e.g. adherence officer or nurse). Each study participant will be assigned to an adherence officer (referred to in this trial as case managers) working in the HIV treatment hospital and participants will be tracked with their mobile phone numbers. The recruitment process is expected to last for 3 months.

### Randomization

The two study sites will be considered as two clusters in the trial, in which patients in each cluster are randomized into treatment and control arm. Randomization will be done with the aid of a research randomizer, a web-based computer random-numbers generator (28). The randomization will be carried out by an independent person who is not part of the research team.

### Outcomes

The primary outcome measure will be the difference between the HIV viral load suppression (≤20 copies/ml) between the baseline, 12 months, and 24 months in both the intervention and the control arms. The secondary outcome measure will include the pill count (it measures adherence), cost-effectiveness, and patient satisfaction. COBAS TaqMan 96 and the Amplilink software will be used to analyze viral load.

### Time frame

April 26^th^, 2021 and it will end on May 10^th^, 2023(see fig 2)

### LABORATORY ANALYSIS

Blood will be collected from the participants (10 mLs) into a vacutainer EDTA-specimen bottle. The EDTA-blood sample will be submitted to the virology laboratory at Nnamdi Azikiwe University Teaching Hospital for routine HIV-1 viral load testing by the CAP/CTM v20. The EDTA-sample tubes will undergo centrifugation at 145g for 25 minutes before the plasma separation into two phases in 2ml cryovials. The sample will be stored at -880C in an ultralow freezer. Manufacturer’s instruction on the Roche CobadAmpliprep/ CobasTaqman (CAP/CTM) commercial kits will be used to measure the viral loads and the plasma HIV-1 RNA levels will be determined as copies/mL

The automatic extraction of the viral RNA will be done and then transferred to the COBAS TaqMan for amplification and detection. The CAP/CTM v2.0 will run along with the quantification range of the CAP/CTM v2.0 from 20 copies to 107copies/mL. Each run will consist of the test samples alongside high positive control, one low positive control, and one negative control from the kits. The result will be validated when the three controls have passed a run. CD4+ count analysis will be performed with the CD4+ easy count kit and the Parteecyflow using the specifications as described by the manufacturer’s manual. The result from the viral load is entered into a case report form (see. Fig5).

### Patient and Public involvement

Patient involvement will be facilitated through contact with people living with HIV/AIDS (PLHA) support groups and HIV/ AIDS focal persons that are based in the study sites. A partnership will be established with three representatives from these groups and the research team. One member will be part of the Project organization, who will advise on study protocol (this includes protocol amendment) and will also participate in explaining the study to the participants in the simplest form, and feedback from the patients will be noted. Two members will be involved as a member of the Project Steering Committee in the study procedures, especially those involving the recruitment and drug delivery process in a manner as to ensure strict confidentiality. One member will also participate in monitoring and evaluation (Data Monitoring Committee) of all the stages in the trial, where he/she will be allowed to comment on and help inform the improvement of the trial at any stage. The public will be involved during the dissemination of the research protocol.

### Data Management

The research assistant is responsible for the collation and processing of the laboratory results, checklist for adherence, and case report form. To ensure confidentiality data will be protected by a password. The research assistant will carry out verification of the forms and capture them on the computer. The chairman of the data management team carries out data verification, data validation, queries data forms that may have issues. The major risk that may occur in this study is the exposure of patient personal information, to curtail this risk patient data will be treated with the utmost confidentiality. HDPs were instructed to seek privacy that is comfortable for the patient and himself before administering the medication and structured interview questions. All data will be collected and processed according to the guidelines on confidentiality established by the National Health Research Ethics Committee (NHREC). Unique identifiers and codes will be used to protect patient data which will be received by the data analyst and the principal investigator. Adverse events (AEs) will be documented and reported to the Data and Safety Monitoring Board and National Agency for Food and Drug Administration (NAFDAC) pharmacovigilance center. In the case of a serious adverse event (SAE), it will be reported to the Nnamdi Azikiwe University teaching hospital for further care, and the patient is withdrawn from the study.

### Statistical analysis

Statistical analysis will be conducted using IBM SPSS for Windows, Version 25 (NY, USA; IBM Corp). The demographic characteristics (as well as outcomes) will be reported as means and standard deviation for both the intervention and control arm. All the primary data will be analyzed using the intention-to-treat principle (to enable us to draw an unbiased conclusion about the effectiveness of our intervention) as well as per-protocol analysis. The latter will help identify how the treatment effect will be under optimal conditions. Logistic regression will be used to predict the factor of events in the primary outcome of this study (i.e., the percentage of participants’ viral load suppression of ≤ 20copies/mls). The balance of covariates will be analyzed by fixed-effects linear regression for continuous confounders (e.g., age) and fixed-effects binomial logistic regression for categorical confounders (e.g., gender). The effectiveness of the treatment will be tested according to patients’ socio-demographics characteristics (age group [40–59 vs. 60–65 years], gender [male vs. female], viral load suppression (≤ 20copies/mls) occupation [working vs. non-working] by conducting a stratified analysis of these covariates. Also, logistic regression will be used to estimate the effect of the treatment while taking into account the secondary outcomes (participants’ viral load suppression of ≤ 20copies/mls vs. adherence, cost-effectiveness, and patient satisfaction). The measure of effect will be the mean difference in outcomes will be derived by subtracting the baseline value from that of the last observation between the intervention and control groups. All statistical tests will be set at one-sided with an alpha level of 0.05.

The Cost-effectiveness of the ‘home-based delivery’ intervention will be assessed. Health resource use will be estimated from the health provider’s perspective i.e. all costs incurred for providing the home-based delivery by a health service provider. Resource use items will include items such as staff time, consumables, cost of financial incentive, and equipment. All patient and family resources will be excluded. An activity-based costing method will be used to measure resource use. Data-capture questionnaires will be developed and sent to all the study personnel to record all their activities or resource use. Resource use items will then be multiplied by unit costs to determine a cost item. The entire cost item will be summed up and divided by the number of participants to determine the cost of the incentive scheme intervention per patient. Cost-effectiveness analysis alongside the randomized trial will be conducted to assess the cost per additional patient achieving viral suppression through the proposed intervention.

The incremental cost-effectiveness ratio (ICER) of ‘Home delivery’ over usual care will be calculated to determine the cost-effectiveness. Patient satisfaction will be evaluated by adopting a validated questionnaire to measure patient satisfaction.

### Program steering committee (PSC)

The program steering committee’s role in this trial is to monitor and evaluate all the steps in research. PSC consists of five members including two patient representatives whose sole responsibility in this trial is to offer surveillance for this research.

### Data monitoring committee (DMC)

The data monitoring committee consists of an independent statistician, nurse, pharmacist, clinician, and patient. The DMC reviews the data and offers advice to other committees in the trial. The DMC will also coordinate the stopping of the trial when the data has been collected. The statistician will conduct a conditional power analysis of interim results or Bayesian predictive probabilities, [29] this will be done to identify if there is a significant difference between treatment groups and it will help the DMC make informed steps on how to end the trial. They will then communicate to the other trial committees to ensure that all aspect of disagreement is addressed before stopping the trial.

### Audit

The last amendment to this protocol was made on 24^th^, April 2021 with a protocol amendment Number: 07. Modifications in the trial will be communicated to the editors for BMJ open journals, trial participants, and ethics committee and trial registries

## DISCUSSION

The findings of this proposed trial should provide evidence on the prospect of delivering ARVs to patients in the comfort of their homes against what is currently obtainable in most health facilities in Nigeria is the constant need to return to a healthcare facility for refill medications rather than a need for clinical monitoring by a nurse or physician (30). It is expected that the Home delivery model can improve patients’ choice for care as well as minimizes interruption to patient’s busy schedule (31,32), and possibly in other sub-Saharan African countries hence improving quality of care and patient satisfaction at the same time suppressing the patient viral load which is crucial for patient survival.

### Ethics and dissemination

Nnamdi Azikiwe University Teaching Hospital Health Research Ethics Committee (NAUTHHREC) approved this study (NAUTH/CS/66/VOL.13/VER III/23/2020/011). The trial will be conducted per the principles outlined in the Declaration of Helsinki. All identifiable data will be handled with the strictest confidentiality. Participants will be informed that they are free to withdraw at any time during any phase of the trial. Dissemination plans involve holding advocacy meetings with stakeholders in HIV care, publishing study outcome in an open-access peer-review journal (with an impact factor), and presentation of the outcome at an international conference.

### Clinical trial registration

This trial is registered in the WHO International Clinical Trials Registry on 8th April 2020 through the WHO International Registry Network with the registration number, PACTR202004535536808.

## Data Availability

The data used and/or analyzed during this study are available from the corresponding author on reasonable request

http://www.ebhc-unizik.com/

## Competing interests

The authors declare that they have no competing interest

### Abbreviations

AIDS: Acquired immunodeficiency syndrome
ARV: Antiretroviral therapy
HIV: Human immunodeficiency virus
CRT: Cluster Randomized trial
SPSS: Statistical Package for the Social sciences
VL: Viral load
WHO: World Health Organization
SDG: Sustainable Developmental Goal
ICER: Incremental cost-effectiveness ratio
NACA: National Agency for Control of AIDS
UK: United Kingdom
SIDSHAS: Strengthening Integrated Delivery of Services on HIV/AIDS
CARC: Community Antiretroviral Refill Club
C-PARP: Community Pharmacy Antiretroviral Refill Programme
UNAIDS: United Nations Programme on HIV and AIDS
HAART: Highly Active Antiretroviral Therapy
HDP: Home delivery Personnel
PLHA: People living with HIV/AIDS

## Acknowledgments

We wish to acknowledge St. Charles Borromeo Specialists Hospital and Iyianu Missionary Hospital for their approval to use their hospital for the study site as well as the provision of the HDPs. Our special appreciation goes to the PLHA support group of St. Charles Borromeo Specialist Hospital and Iyianu Missionary Hospital for their support.

## Author contributions

NA, OE, and SN designed the trial. NA drafted the first protocol. All authors participated in reviewing the protocol. NA submitted the protocol for ethical clearance. All authors read and approved the final version of the protocol

---

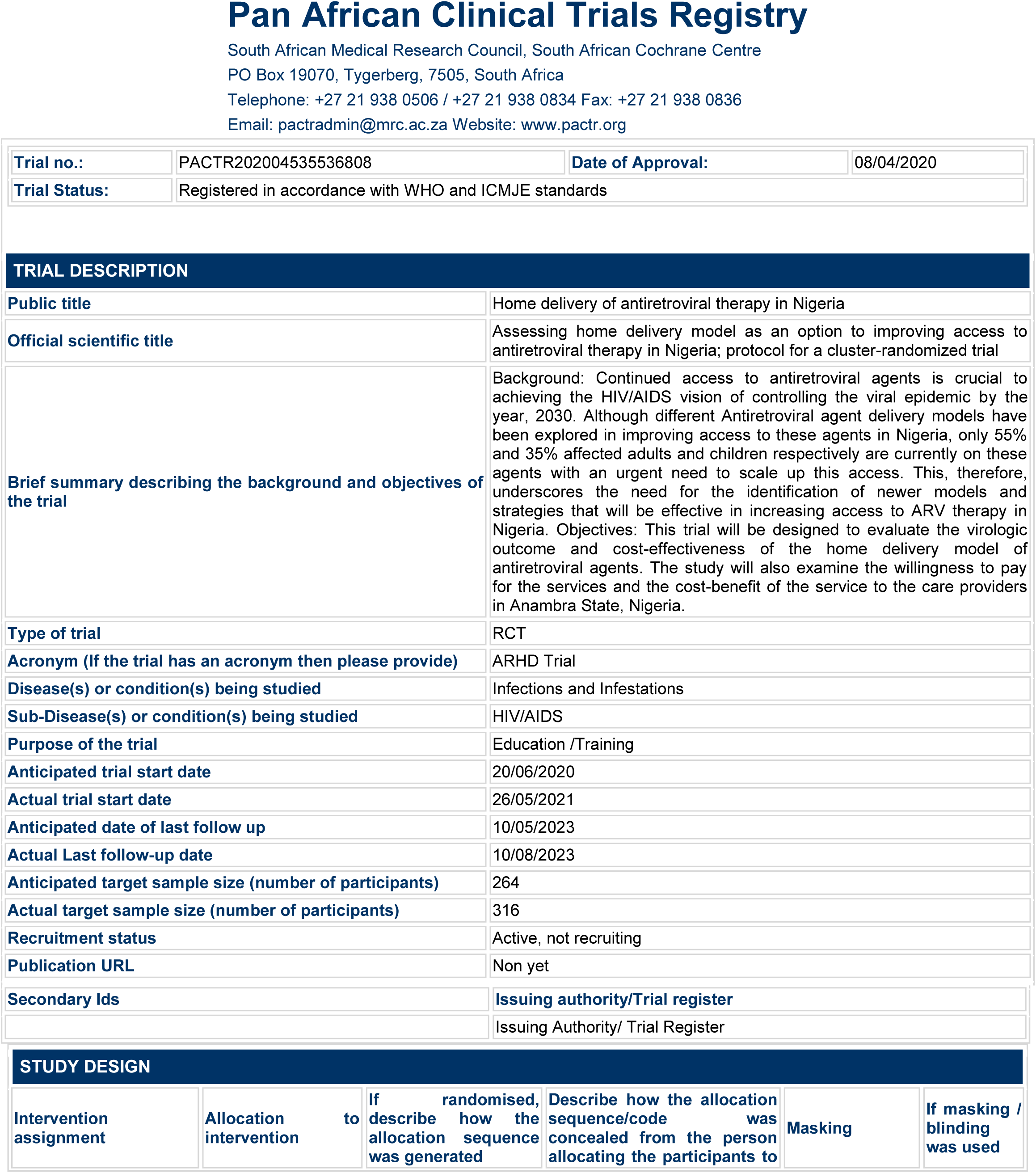

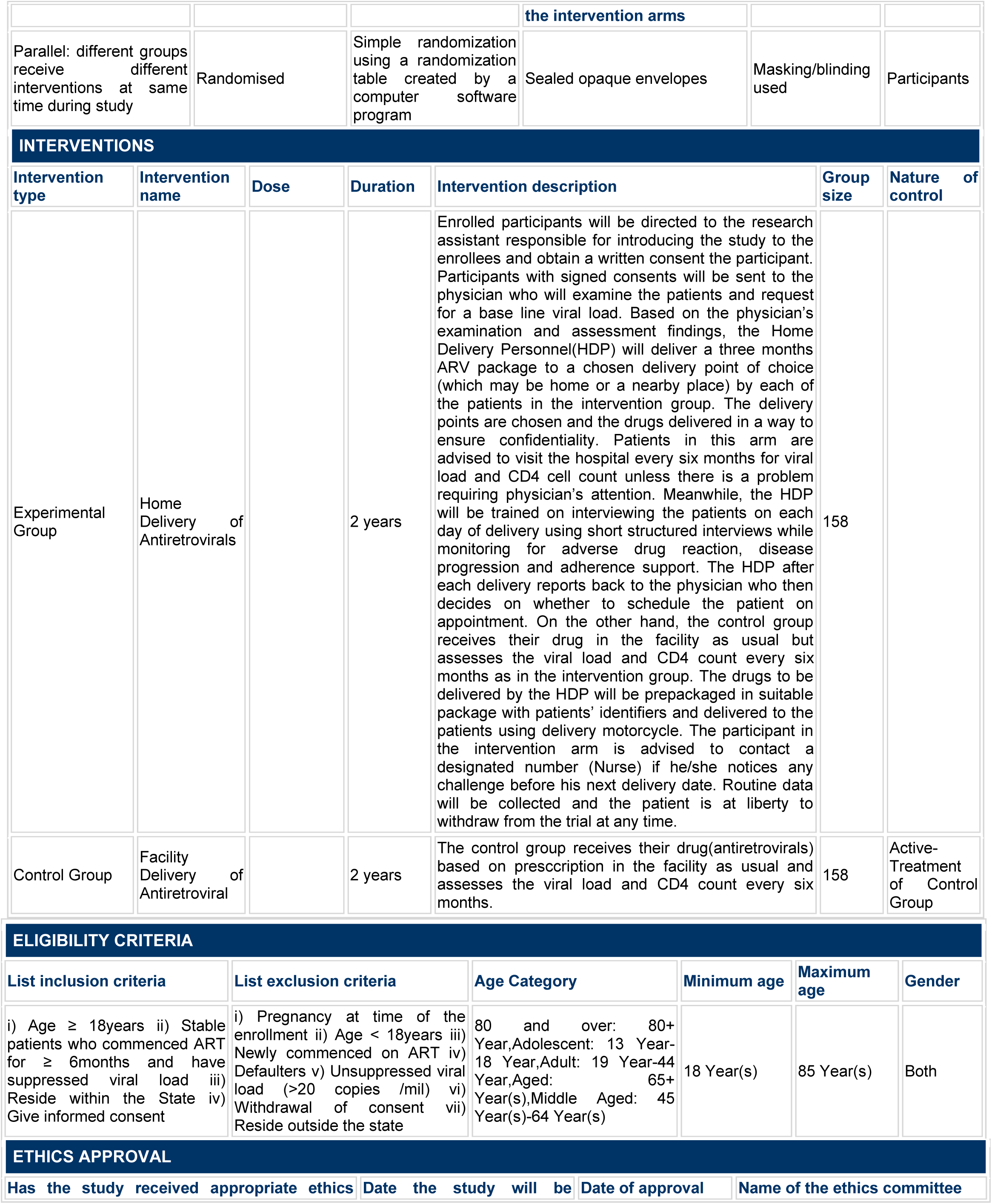

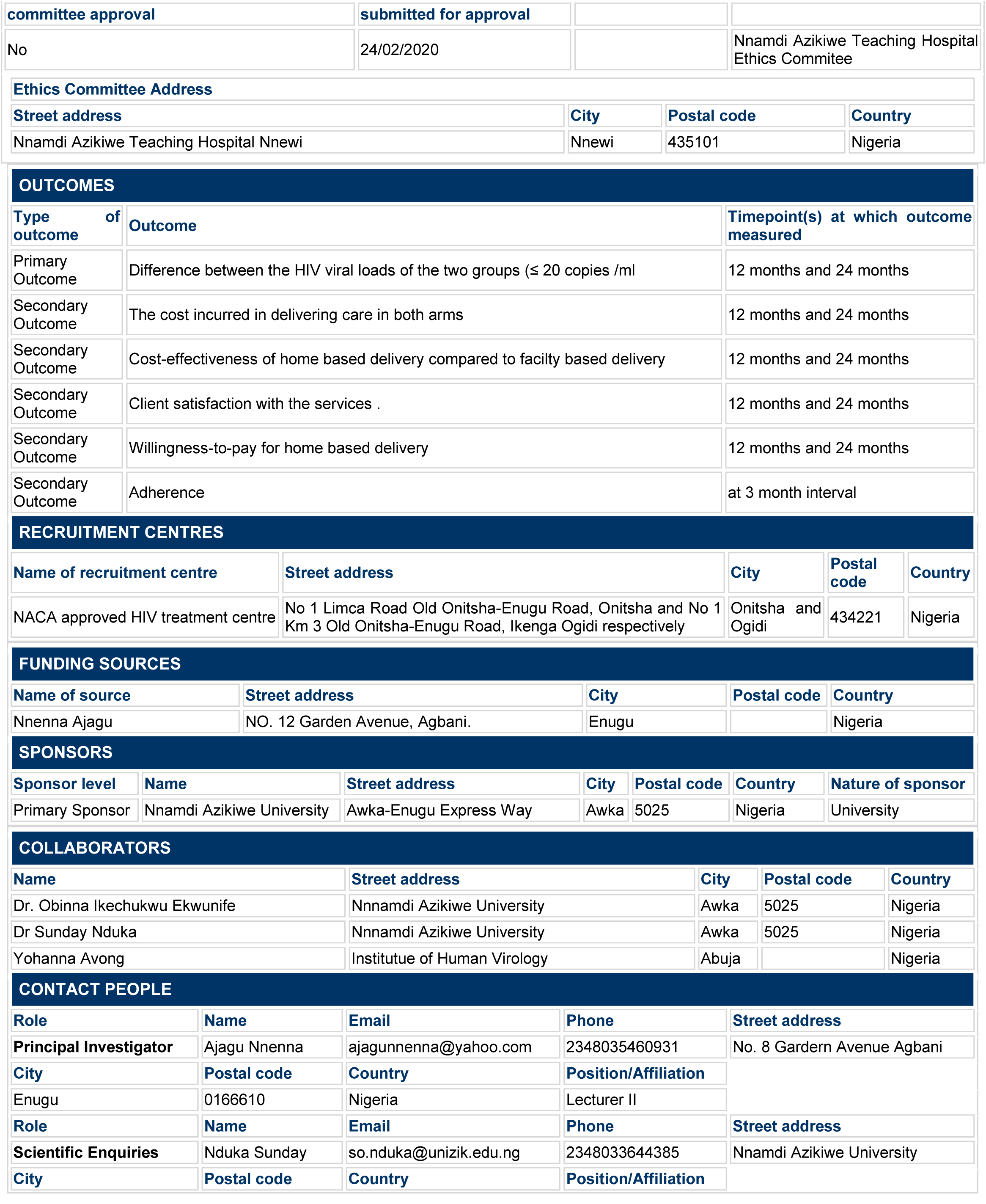

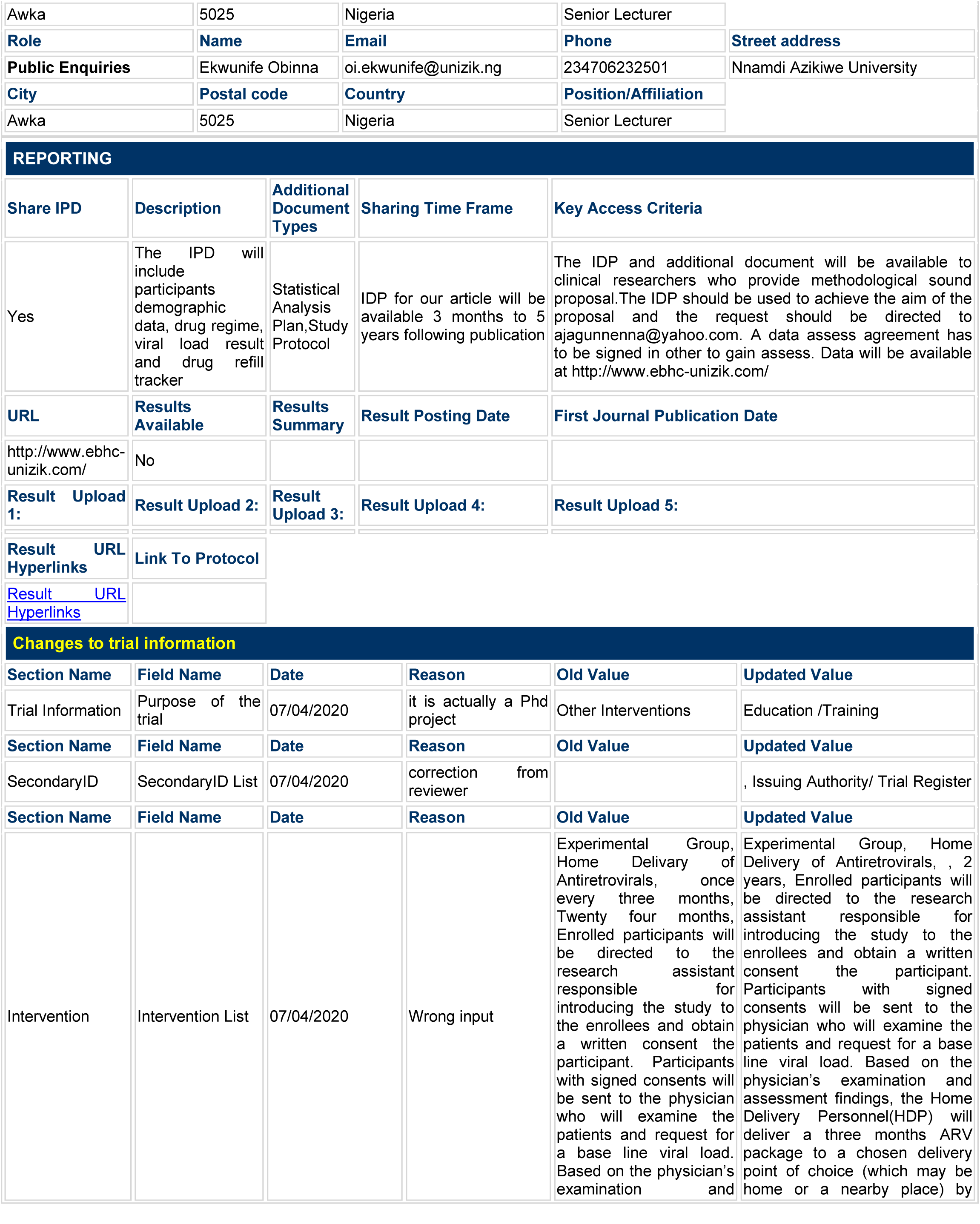

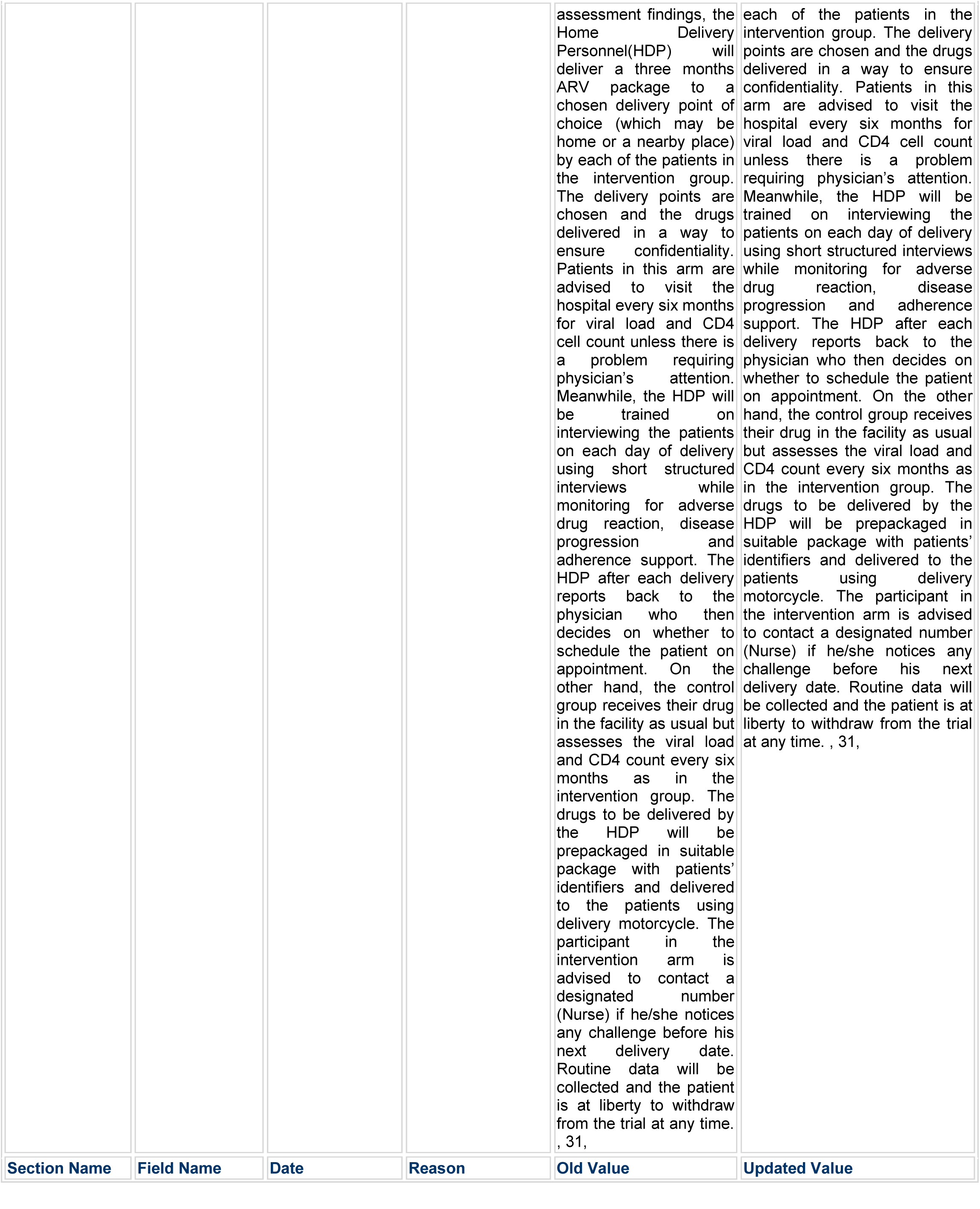

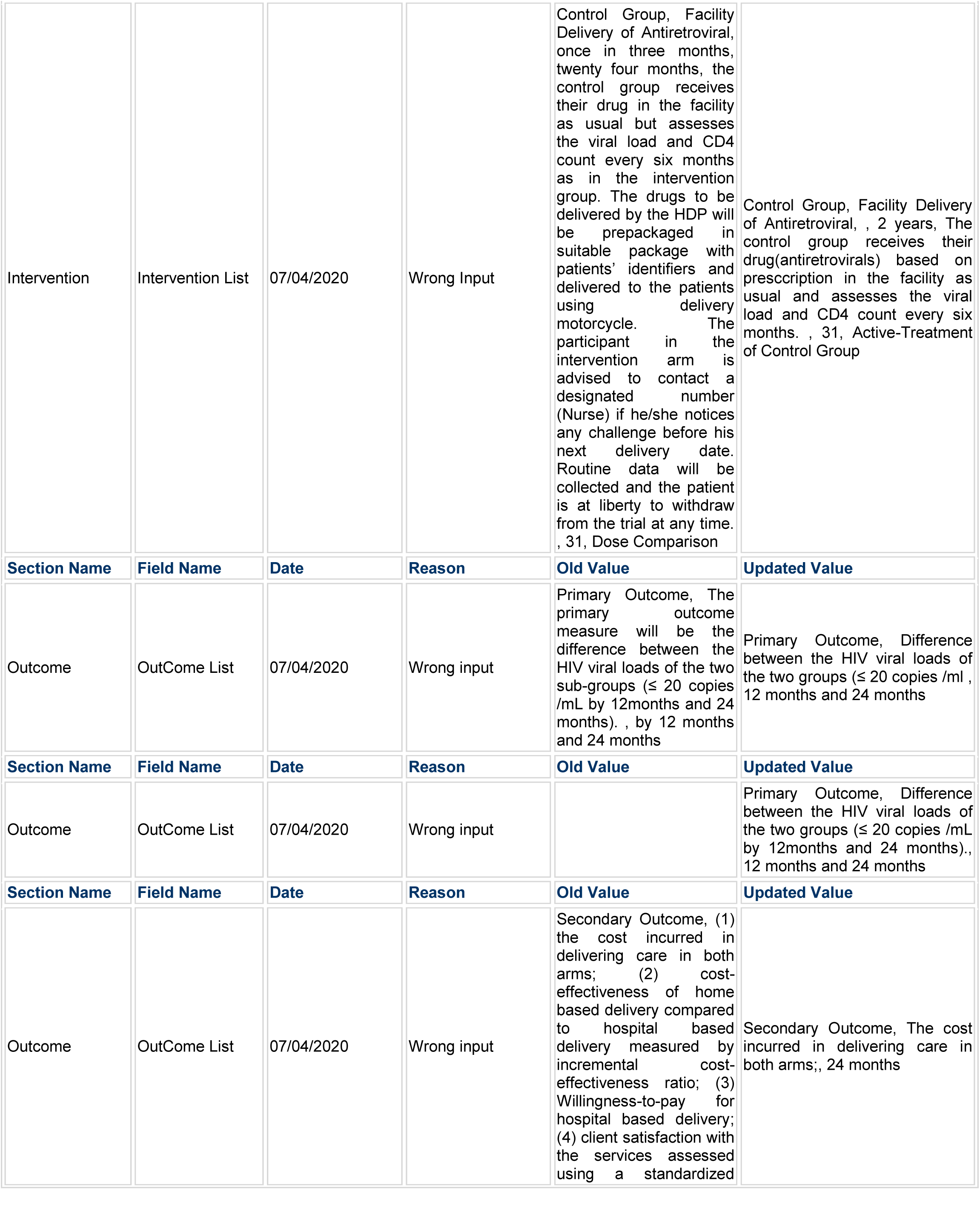

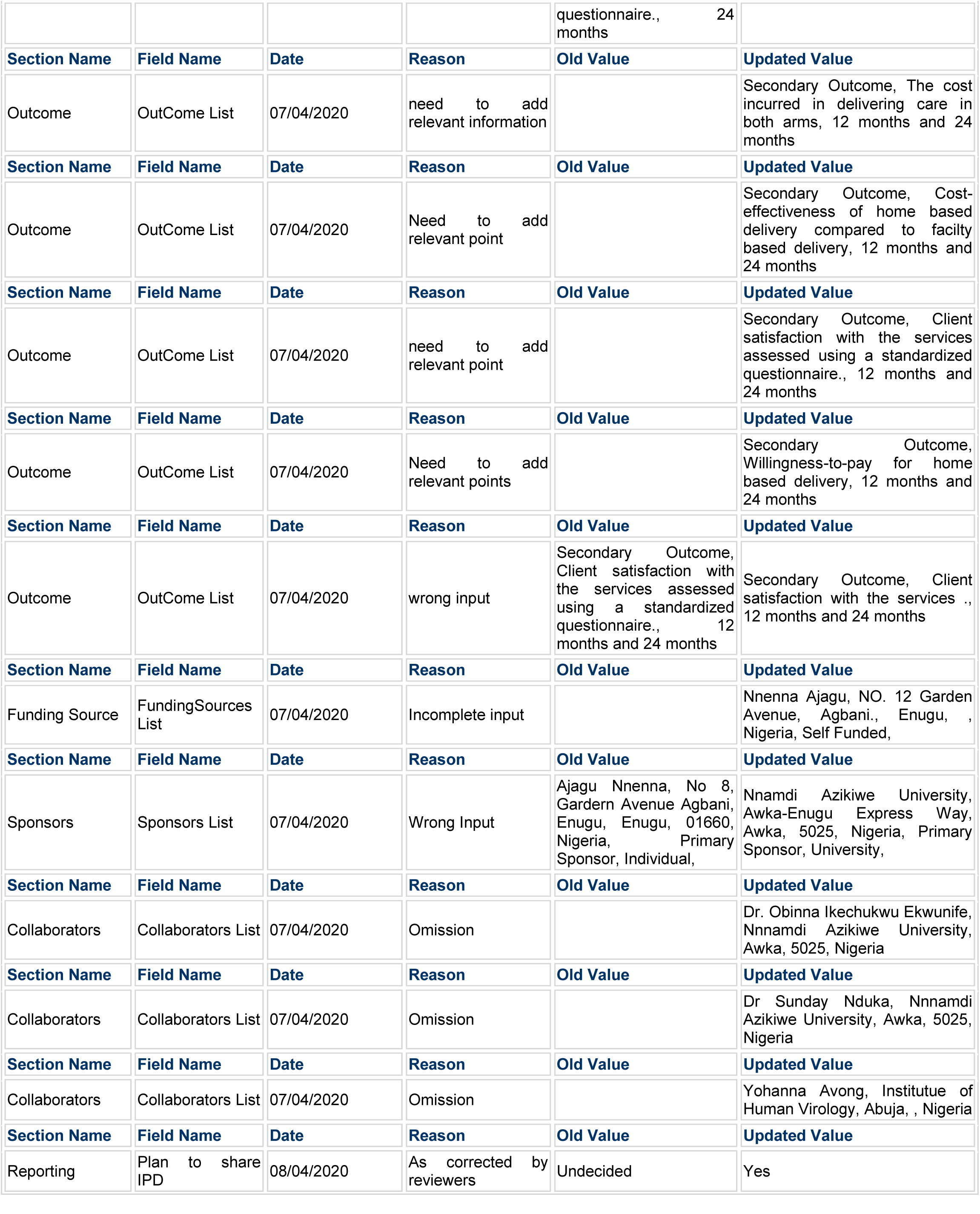

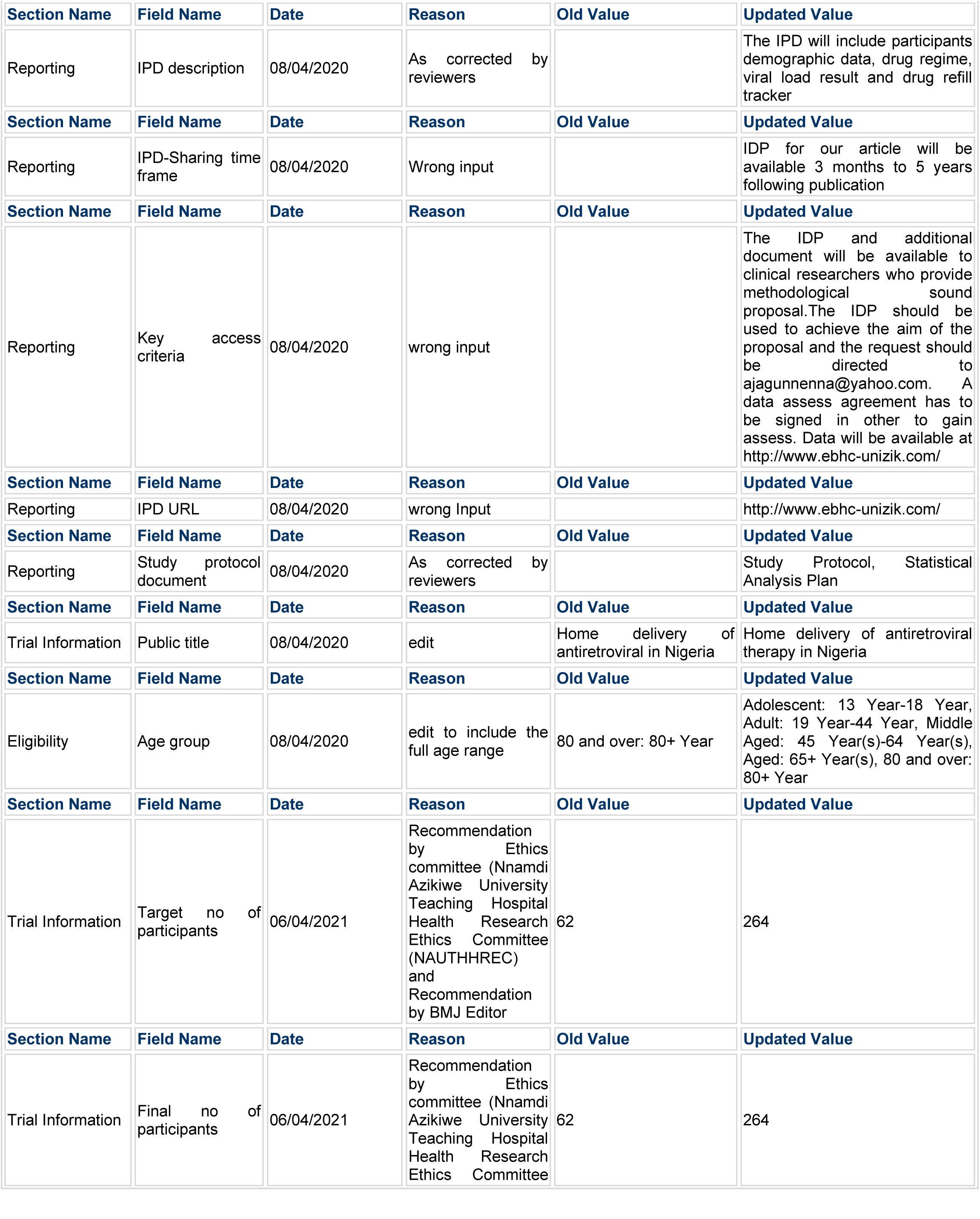

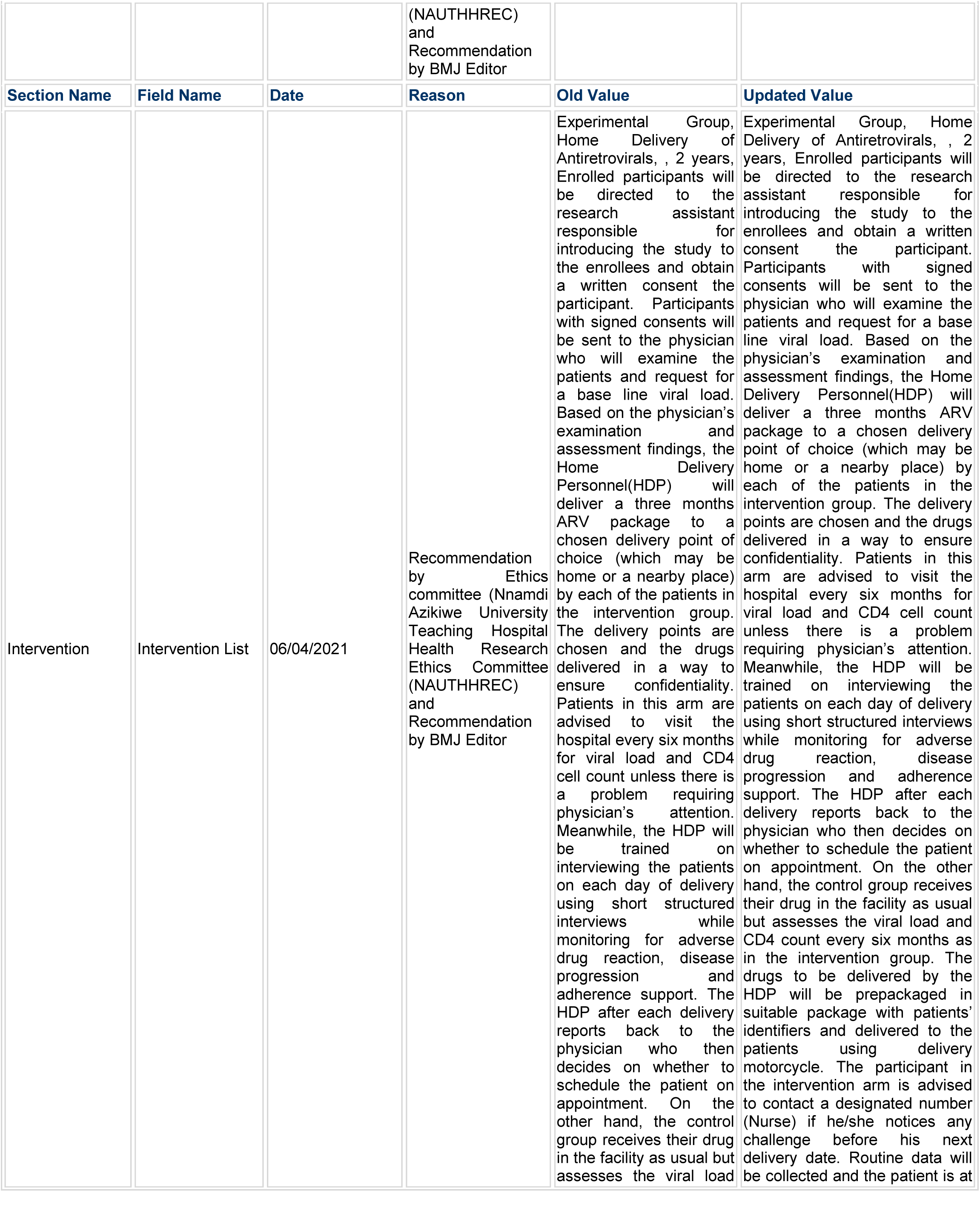

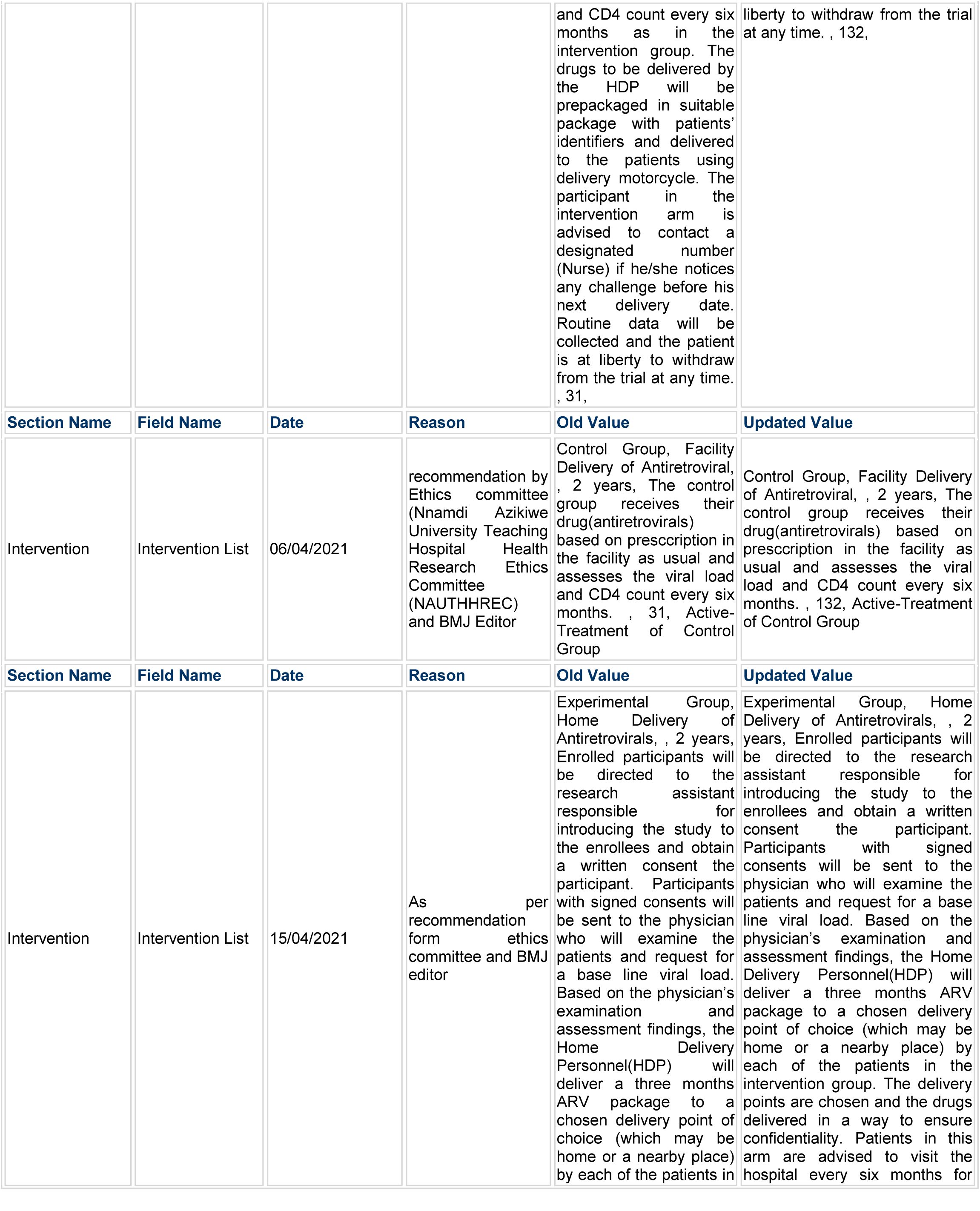

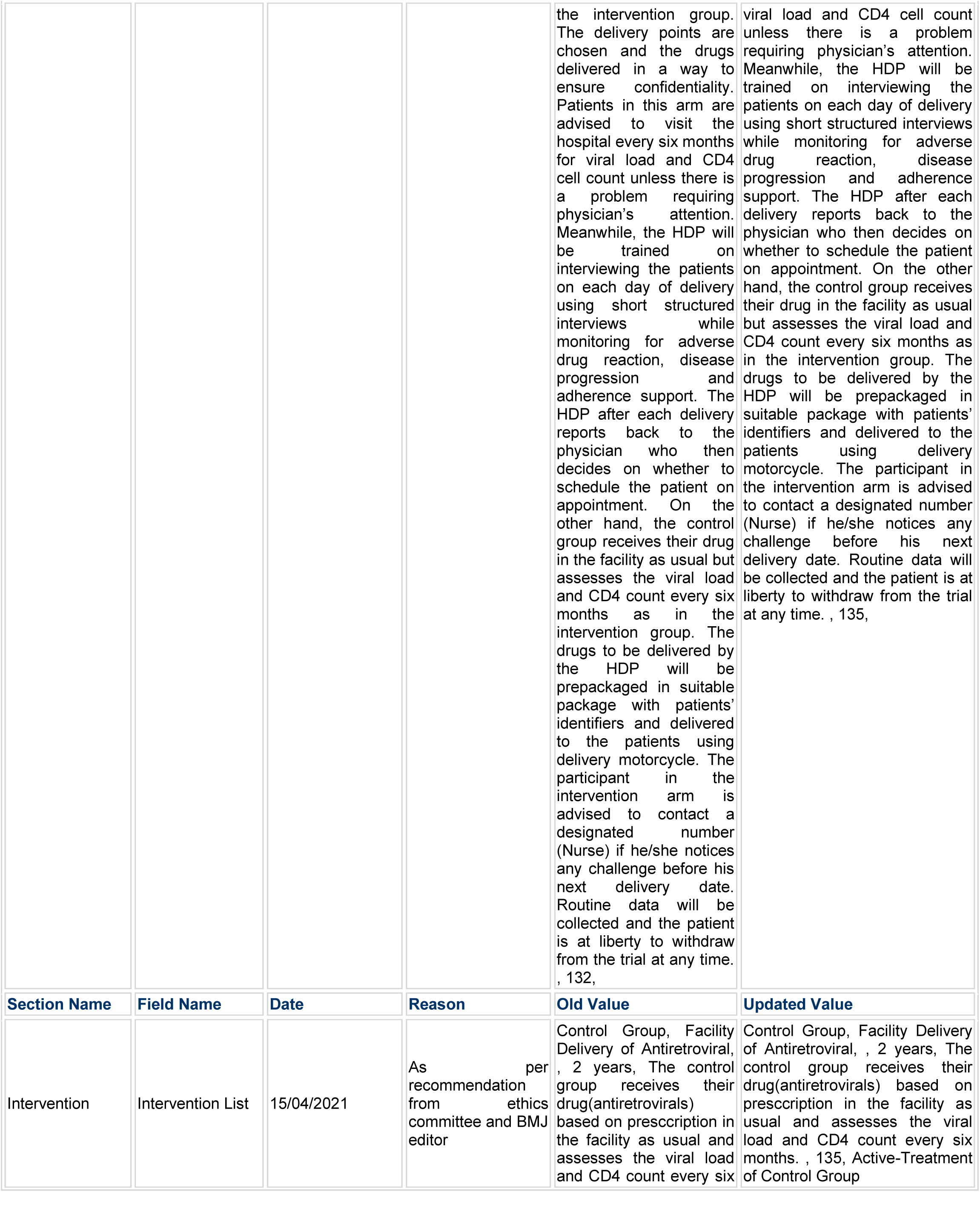

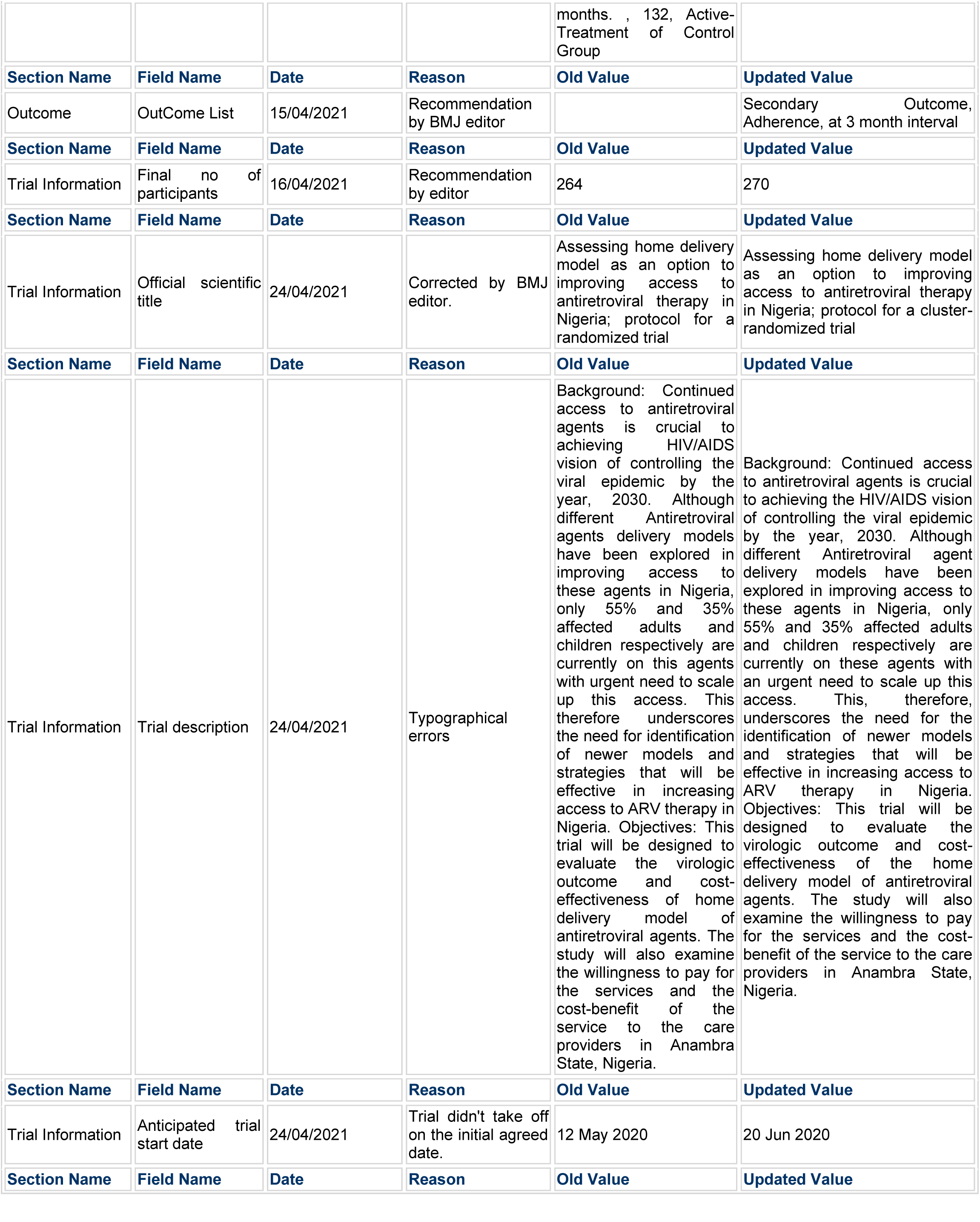

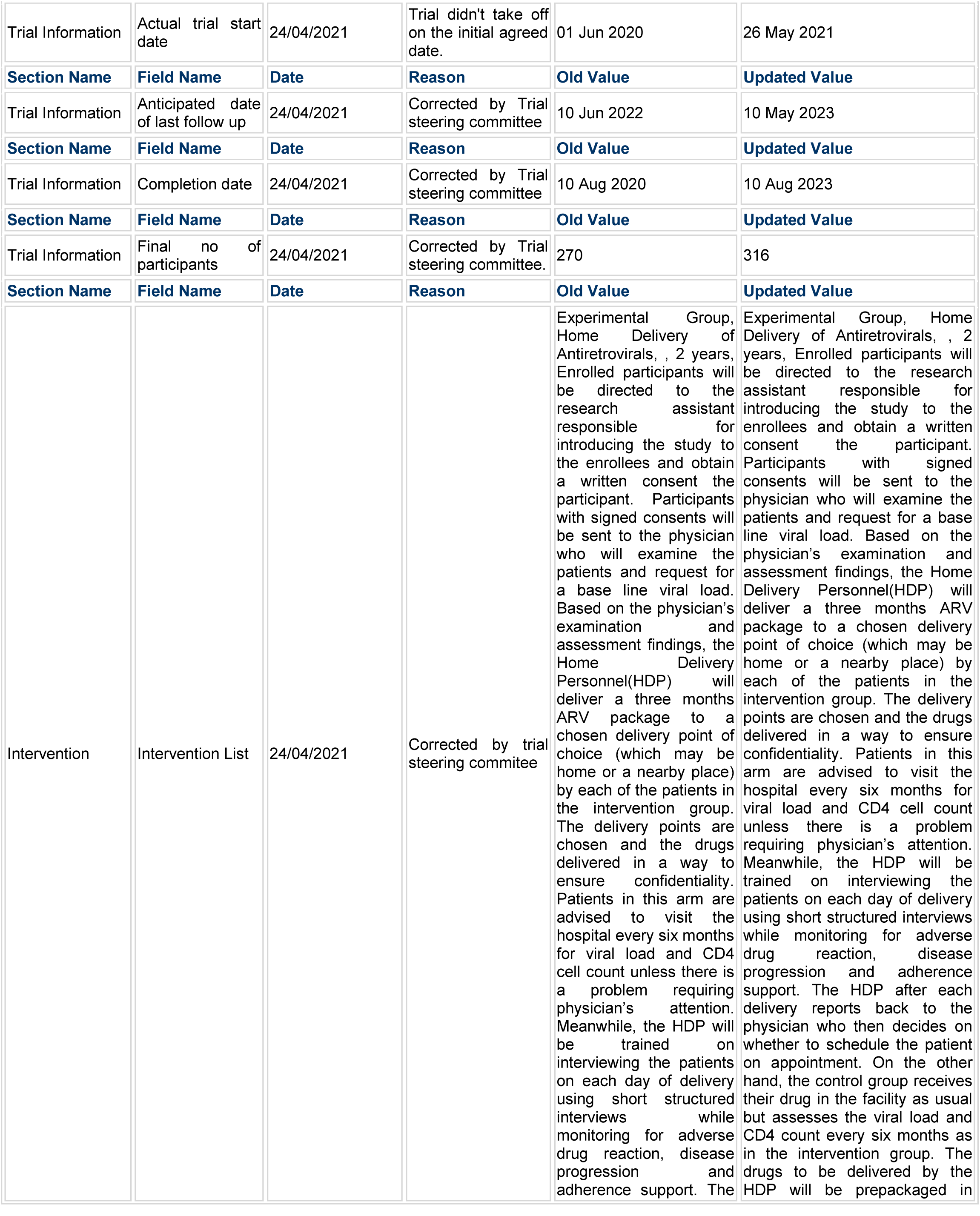

|  |  |  |  | HDP after each delivery reports back to the physician who then decides on whether to schedule the patient on appointment. On the other hand, the control group receives their drug in the facility as usual but assesses the viral load and CD4 count every six months as in the intervention group. The drugs to be delivered by the HDP will be prepackaged in suitable package with patients' identifiers and delivered to the patients using delivery motorcycle. The participant in the intervention arm is advised to contact a designated number (Nurse) if he/she notices any challenge before his next delivery date. Routine data will be collected and the patient is at liberty to withdraw from the trial at any time. , 135, | suitable package with patients' identifiers and delivered to the patients using delivery motorcycle. The participant in the intervention arm is advised to contact a designated number (Nurse) if he/she notices any challenge before his next delivery date. Routine data will be collected and the patient is at liberty to withdraw from the trial at any time. , 158, |
| --- | --- | --- | --- | --- | --- |
| Section Name | Field Name | Date | Reason | Old Value | Updated Value |
| Intervention | Intervention List | 24/04/2021 | Recommendation by trial steering committee | Control Group, Facility Delivery of Antiretroviral, , 2 years, The control group receives their drug(antiretrovirals) based on prescription in the facility as usual and assesses the viral load and CD4 count every six months. , 135, Active-Treatment of Control Group | Control Group, Facility Delivery of Antiretroviral, , 2 years, The control group receives their drug(antiretrovirals) based on prescription in the facility as usual and assesses the viral load and CD4 count every six months. , 158, Active-Treatment of Control Group |

